# Leveraging Large Language Models and Patient Portal Messages for Early Identification of Depression

**DOI:** 10.1101/2025.08.15.25333781

**Authors:** Jiyeong Kim, John Torous, Julia Adler-Milstein, Peter J. van Roessel, Fatima Rodriguez, Christopher Sharp, Michael A. Pfeffer, Jonathan H. Chen, Stephen P. Ma, Carolyn I. Rodriguez, Eleni Linos

**Author notes:** **Correspondence to:** Jiyeong Kim, PhD Stanford Center for Digital Health School of Medicine, Stanford University 3180 Porter Dr, Palo Alto, CA 94304, USA. Contributed equally (Stephen P. Ma, Carolyn I. Rodriguez, Eleni Linos).

## Abstract

**Importance:** Large language model (LLM)-assisted early warning system may help overcome existing barriers to timely depression diagnosis in patients with cardiovascular disease (CVD). This novel application of LLMs to screen patient messages could be applied to other chronic diseases, facilitating automated symptom-driven diagnoses and interventions.

**Objective:** To prospectively simulate the impact (change in time to diagnosis) of population mental health screening using LLMs by screening patient portal messages, and measure LLMs’ accuracy to identify individuals at high risk for depression diagnosis in patients with CVD.

**Design:** Prospective cohort study

**Setting:** Electronic health records from an academic hospital (Stanford Health Care)

**Participants:** Individuals with CVD diagnosed 2014-2024, subsequently diagnosed with depression

**Intervention/Exposure:** LLMs (Llama 3.1 8B, July 2024 version, Meta LLC, and MedGemma 4B, July 2025 version, Google DeepMind, LLC) to identify individuals with depression

**Main outcome:** Accuracy of LLMs in sensitivity for completeness to capture positive cases and positive predictive value (PPV) for correctness to capture positive cases, and changes in time to depression diagnosis

**Results:** We identified 115,156 patients with CVD, and 23.1% (N = 26,578/115,156) of those had co-morbid depression. We included individuals (n=2,314) who sent at least one message between CVD and depression diagnoses. Participants were mostly 65 years and older (n = 1,718/2,314, 74.2%), or of non-Hispanic ethnicity (N = 2,078/2,314, 89.9%), or of the White race (N = 1,506/2,314, 66.1%), but sex was balanced (females, N = 1,197/2,314, 51.7%).

PPV was 51.2% [95% CI: 47.5-54.5%] Llama 3.1 8B, and sensitivity was 83.6% [81.1-85.9] Llama 3.1 8B and 71.0% [67.0-75.3] MedGemma 4B. On average, the LLM (Llama 3.1 8B) detected depression 660 days earlier than the first charted diagnosis over a 1,746-day assessment period, a typical timeline from CVD to depression diagnoses in our cohort.

**Conclusion/Relevance:** LLMs identified individuals with depression significantly earlier than official diagnosis among patients with CVD, relying solely on longitudinal patient messages without additional medical information, with high sensitivity and Patient Health Questionnaire-9 comparable PPV. This novel approach is applicable to various diagnoses.

**Key points:** *Questions:* Among those with cardiovascular disease, can large language models (LLMs) identify individuals with depression earlier than the first charted diagnosis via patient messages, and how much sooner and accurately? How frequently are depression symptoms detected before diagnosis?

*Findings:* LLMs identified individuals with depression 660 days earlier than the official diagnosis, with promising sensitivity (83%) and Patient Health Questionnaire-9 comparable PPV (51%)

*Meaning:* LLMs applied to patient messages showed strong potential for early diagnosis as a screening assistant, which is a novel use for this data source that could be expanded to other diagnoses.

## Introduction

Co-morbid depression among individuals with cardiovascular disease (CVD) is a significant contributor to the global burden of disease, leading to premature death and disability.^1–3^ Individuals with CVD have a 2-3 times higher prevalence of depression compared to those without CVD.^4–6^ CVD and depression are intimately related: individuals with CVD are more likely to develop depression, and depression is a risk factor for developing CVD.^7^ A plausible link between CVD and depression may involve common physiological disruptions, such as increased inflammatory markers, endothelial dysfunction, and hyperactive platelets.^5,8,9^ Untreated depression exacerbates CVD progression, complicating disease management and deteriorating health outcomes and quality of life for this vulnerable population.^2,7,10^

Timely diagnosis and treatment initiation for depression are critical, yet diagnosis is often delayed due to a number of barriers. Patients may not be aware of their symptoms or may lack motivation to seek care, as both medical and mental disorders can cause similar somatic symptoms like fatigue, reduced appetite, and changes in sleep, limiting the effectiveness of self-screening.^2^ In non-psychiatry clinics, particularly primary care settings, clinicians may struggle to diagnose co-morbid depression in a timely manner in patients with chronic medical conditions, including CVD.^11^ There are multiple contributing factors, including less practice identifying subtle symptoms of depression, a focus on the many other medical concerns that need to be addressed during typical primary care visits, and limited time allocated for each patient.^11–14^

Patient portal messaging has become an essential communication channel that allows patients to securely share their clinical symptoms and asynchronously address health issues with healthcare professionals.^15^ Longitudinally compiled patient-reported health information in such messages can potentially be a valuable source for detecting chronic conditions that develop over time, but are not currently utilized for population health screening, given the amount of manual labor that would be required. Large language models (LLMs) have emerged as transformative tools in mental health,^16–18^ with the ability to detect symptoms of depression and anxiety from patient-generated text data and from patient messages of those with diabetes.^19,20^ This suggests the potential of LLMs to identify depression through high-throughput automated screening of patient portal messages before clinicians recognize clinically meaningful symptoms, even without patients’ full awareness of their own symptoms.

However, this potential application of LLMs remains underexplored despite its potential to enable early diagnosis of depression in the high-risk population of patients with CVD. Here, we prospectively simulate the impact of population health screening using LLMs by screening patient portal messages for the identification of patients at high risk for a new diagnosis of depression in the cohort of patients with a recent diagnosis of CVD. In addition to classic performance metrics, we also explicitly measured the potential impact on patients as the change in time to diagnosis.

## Methods

### Data source

We obtained de-identified secure patient messages from individuals with CVD (ICD-10 codes of I20, I21, I25, I42, I49, I50, and I63) between 01/2014 and 07/2024 at a large academic medical center (Stanford Health Care, SHC). We focused on patient medical advice requests (PMARs), routed to internal medicine, family medicine, or primary care, excluding scheduling, medication refill requests, and insurance-related inquiries. We performed data post-processing for additional data security, and all the analyses were done in a HIPAA-compliant analytics environment. The Stanford University Institutional Review Board approved this study.

### Data preparation

To identify case individuals, those with CVD and co-morbid depression, we used the ICD-10 code of F32 (a single episode) documented within the patient’s electronic health record (EHR). We intentionally used F32, not including F33 (recurrent episodes), to identify people who developed depression only after CVD diagnosis. Among individuals with both CVD and depression, those who were diagnosed with CVD first, then later diagnosed with depression, and sent at least one secure message between CVD and depression diagnoses were included as cases to be eligible for LLM assessment. Then, we extracted the message conversations of the eligible case patients that occurred between the two diagnoses. We included the messages from patients only, while excluding those from healthcare professionals. To prepare them for two different analyses, we cleaned the message data by removing empty lines, subject lines, and duplicates, and then ordered them chronologically.

To obtain the usual diagnostic timeline of patients with CVD to be diagnosed with depression, longitudinally following them in the EHR for 10 years (2014-2024), we computed the time difference from the date of CVD diagnosis to the date of depression diagnosis, using the first date each ICD-10 code was documented in the patient’s EHR. To understand the typical timeline between CVD and depression diagnoses in our cohort, we calculated the mean (m) time difference with standard deviation (SD).

To identify control individuals, those with CVD but without co-morbid depression, we performed matched sampling using case individuals’ sociodemographic characteristics such as (18-34 years, 35-49 years, 50-64 years, 65-74 years, and 75 and older), race (Asian/Pacific Islander, Black/African American, Native American/Other, and White), ethnicity (Hispanic and non-Hispanic), and sex (female and male) to minimize potential selection bias. To collect their secure messages sent for the same duration as in our case population, we used the mean time difference between CVD diagnosis and depression diagnosis as calculated above.

### LLMs used in this study

For all experiments in this study, we used two open-source small LLMs that can be further optimized for this use case (Llama 3.1 8B, July 2024 version, Meta LLC, and MedGemma 4B, July 2025 version, Google DeepMind, LLC). We chose Llama 3.1 8B because it demonstrated exceptional zero-shot performance for detecting depression symptoms from patients’ text messages.^20^ We also evaluated MedGemma 4B, which is a recently developed powerful and cost-efficient model fine-tuned specifically for medical text comprehension.^21^

### Threshold determination

The messages of those with CVD and depression, chronologically ordered and noted by the message received date and time, were first evaluated by LLMs for the presence of depression symptoms. Each individual message was classified as “0” for no symptoms or “1” for symptoms. The evaluated messages were then organized under the patient.

Next, to determine the number of symptoms that should represent a true depression diagnosis, we computed the frequency of depression symptoms detected from secure patient messages per patient until they were formally diagnosed. We used the median and median absolute deviation (MAD) because the data were not normally distributed.

### Prospective Simulation of Population Screening

Then, we simulated whether the LLM could identify individuals with depression among those with CVD from longitudinally compiled messages. For efficiency, we used a random sub-sample of 1,000 patients; 500 were cases labeled as ‘1’, representing those with CVD and depression.

Another 500 were controls labeled as ‘0’, representing those with CVD but without depression. The average number of compiled messages per patient was computed to provide an overview of the descriptive difference between the two groups. We compared the performance of LLM screening using a variety of thresholds (e.g. threshold=3, “Classify the following message as 1 if the patient shows depressive symptoms at least three times, otherwise 0”), including the median number of messages marked as “1” for symptoms identified above, against the baseline performance of LLMs asked to identify patients with depression without a specific pre-determined threshold (”Classify the following message as 1 if the patient shows depressive symptoms, otherwise 0”). A full prompt is available in eMethods 1. Screening performance was calculated using classic metrics, including sensitivity to show the completeness of capturing positive cases and positive predictive value (PPV) to show the correctness of capturing positive cases with 95% confidence intervals derived via bootstrapping (n=1,000).

### Changes in time to diagnosis

Finally, for the case cohort, we marked the date of the message sent for which the median frequency threshold was reached as the time point of an LLM’s diagnosis. Then, we measured the time difference between this time point and the formal diagnosis of depression to assess the impact of LLM screening on the timeliness of diagnosis.

## Results

### Descriptive data flow

We identified 115,156 patients with CVD in the past 10 years, of whom, 23.1% (N = 26,578/115,156) had co-morbid depression. Among these, 48.5% did not have depression at the time of CVD diagnosis and were diagnosed with depression subsequently (N = 12,899/26,578) (**Figure 1**). The number of secure messages sent between CVD and depression diagnoses was 58,166 from 2,314 individuals.

**Figure 1.**
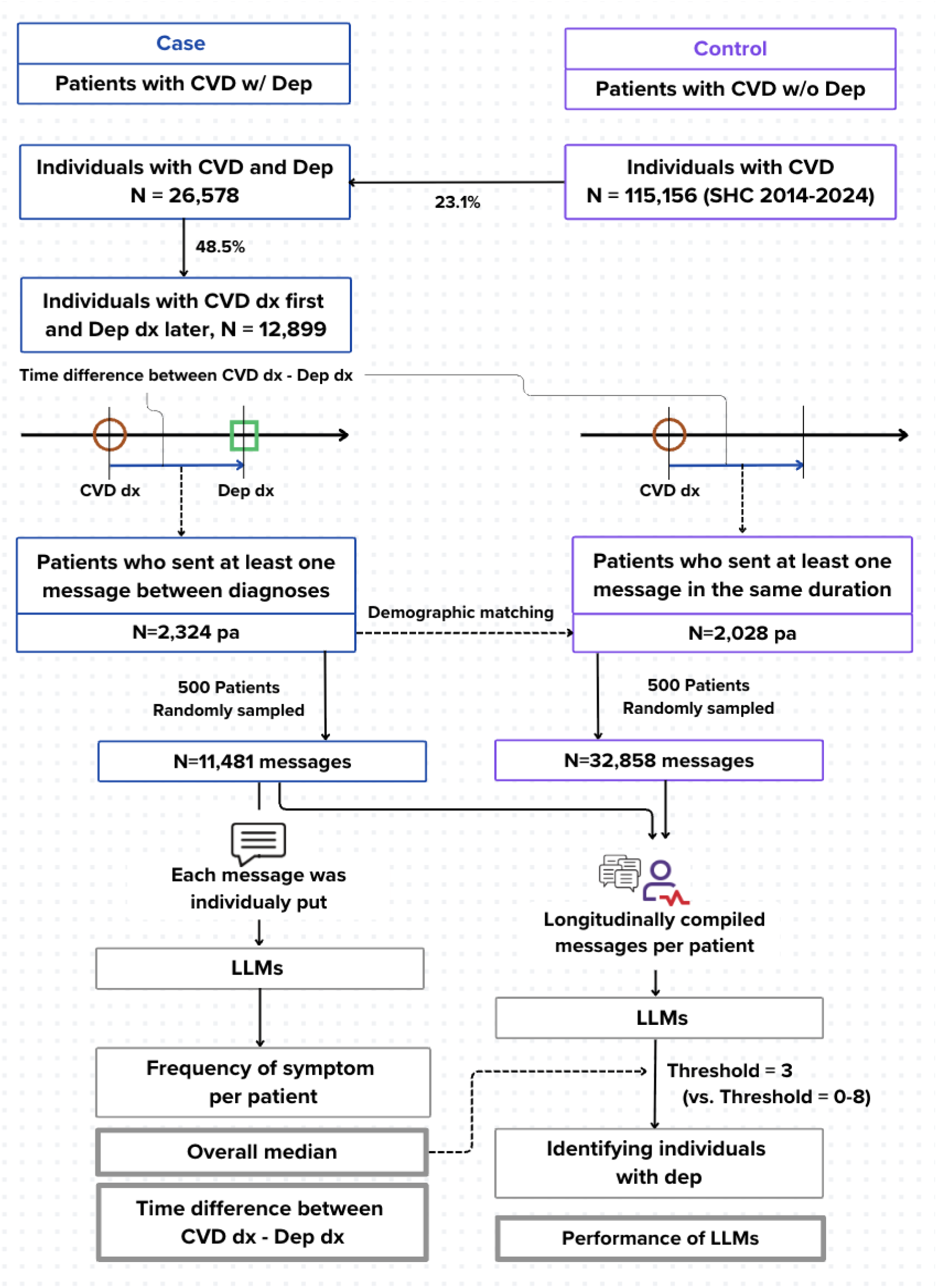
Study design and data flow. a. Age, race, ethnicity, and sex were matched; b. LLMs: Llama 3.1 8B and MedGemma 4B; c. Threshold: the frequency of depression symptom presentations for LLMs to identify individuals with depression; Abbreviations: CVD (Cardiovascular disease), LLM (Large language model), Dep (Depression), dx (Diagnosis), SHC (Stanford Health Care)

When the LLM classified each message for the presence of depression symptoms, the median frequency was 3.0 (MAD: 2.0) across 58,166 messages from 2,314 patients with CVD and depression among those who sent at least one message (Llama 3.1 8B).

### Sociodemographic characteristics

**Table 1** shows sociodemographic characteristics of the included individuals. Patients with CVD and co-morbid depression were mostly 65 years and older (n = 1,718/2,314, 74.2%), or of non-Hispanic ethnicity (N = 2,078/2,314, 89.9%). Approximately two-thirds were of the White race (N = 1,506/2,314, 66.1%) while Asian/Pacific Islander/Native American/Other races were one-third (N = 654/2,314, 29.0%) and 5% (N = 120/2,314) were Black/African American. Females (N = 1,197/2,314, 51.7%) were slightly more than males (N = 1,117/2,314, 48.3%).

**Table 1.** Sociodemographic characteristics of patients with cardiovascular disease.

|  | **CASE:**<br>**Patients with CVD and co-morbid depression**<br>**(N=2,341)** |  | **CONTROL:**<br>**Patients with CVD and without co-morbid depression**<br>**(N=2,028)** |  |
| --- | --- | --- | --- | --- |
|  | Frequency (N) | (%) | Frequency (N) | (%) |
| **Age** |  |  |  |  |
| 18-34 | 37 | 1.6 | 32 | 1.6 |
| 35-49 | 146 | 6.3 | 131 | 6.5 |
| 50-64 | 413 | 17.8 | 376 | 18.5 |
| 65-74 | 540 | 23.3 | 494 | 24.4 |
| 75+ | 1178 | 50.9 | 995 | 49.1 |
| **Race** |  |  |  |  |
| Asian/Pacific Islander | 417 | 18.3 | 377 | 18.8 |
| Black | 120 | 5.3 | 98 | 4.9 |
| White | 1506 | 66.1 | 1322 | 66.1 |
| Native American/Other | 237 | 10.4 | 204 | 10.2 |
| Unknown | 34 | 1.5 | 27 | 1.3 |
| **Ethnicity** |  |  |  |  |
| Hispanic/Latino | 175 | 7.6 | 151 | 7.4 |
| Non-Hispanic | 2078 | 89.8 | 1824 | 89.9 |
| Unknown | 61 | 2.6 | 53 | 2.6 |
| **Sex** |  |  |  |  |
| Female | 1197 | 51.7 | 1063 | 52.4 |
| Male | 1117 | 48.3 | 965 | 47.6 |

Case and Control: Individuals were identified by ICD-10 codes for cardiovascular disease and depression (Stanford Health Care, 2014-2024); Abbreviations: CVD (Cardiovascular disease)

### Prospective simulation results

When applying the median frequency threshold of three symptom presentations, the LLM showed highest PPV (51.2% [95% CI: 47.5-54.5%] Llama 3.1 8B), while sensitivity was highest (83.6% [81.1-85.9] Llama 3.1 8B; 71.0% [67.0-75.3] MedGemma 4B) with threshold of two symptoms (**Figure 2**). Performances largely varied by threshold in MedGemma 4B, including sensitivity from 20.2% (threshold = 0) to 71.0% (threshold = 3), while performances less varied by threshold in Llama 3.1 8B, including sensitivity from 67.4% (threshold = 0) to 83.6% (threshold = 3). The baseline performance (without the threshold) was poorest (PPV 47.1% [43.4-50.8] and sensitivity 67.4% [64.5-70.2] in Llama 3.1 8B; PPV 35.3% [29.6-40.8] and sensitivity 20.2% [16.9-23.9] in MedGemma 4B) than those with the threshold.

**Figure 2.**
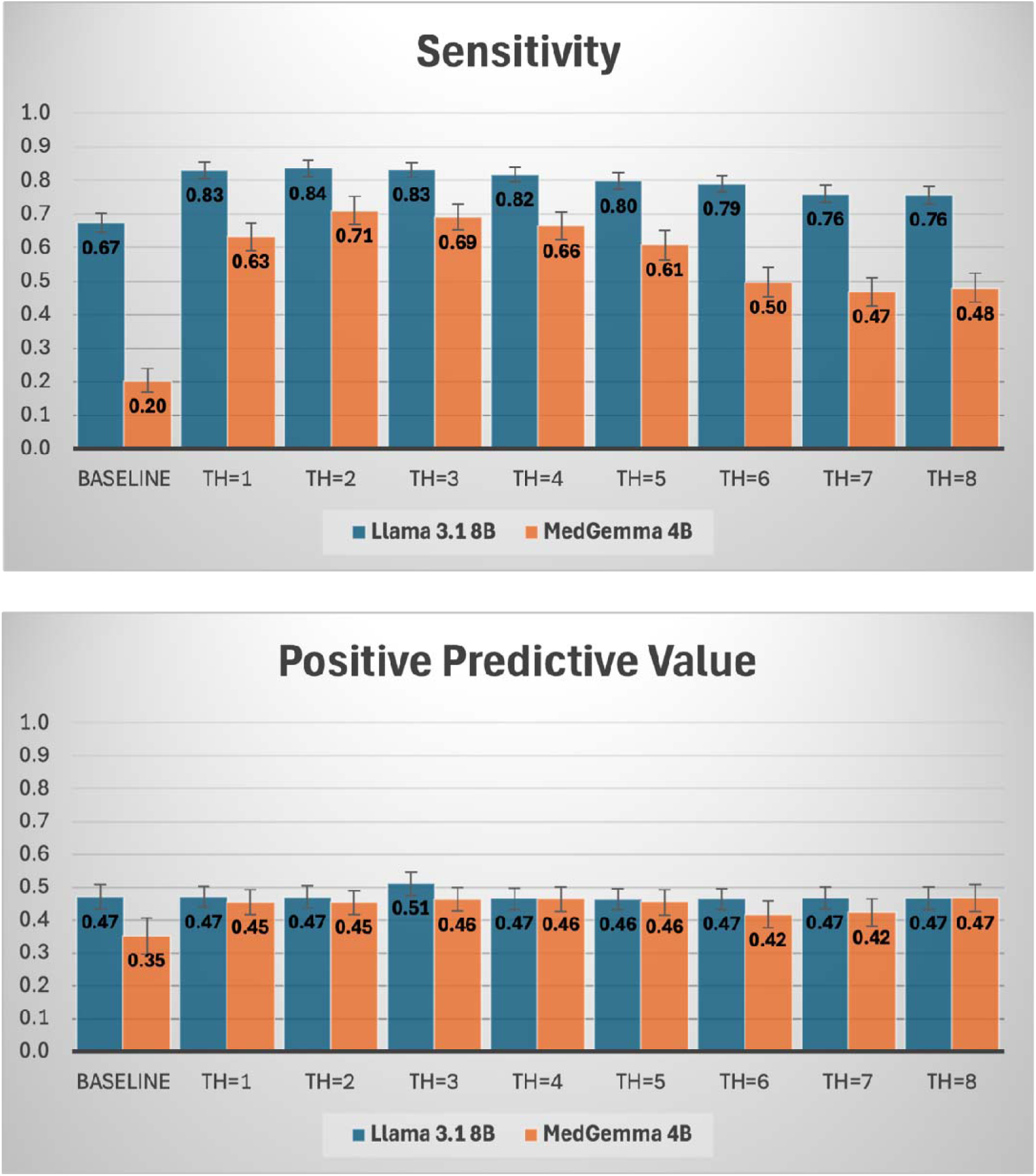
**Performance of LLMs to identify individuals with depression among those with CVD from patient messages by threshold** a. Baseline means that there was no specific threshold to identify individuals with depression (LLMs determined). b. TH: threshold (e.g. TH=1 means that LLMs identified individuals who showed at least one depression symptom) c. Reference standards for computing sensitivity and positive predictive value: cases (individuals with CVD and co-morbid depression) were “1” and controls (individuals with CVD but no co-morbid depression) were “0”

### Timeliness of LLMs’ depression diagnosis

In this study, patients with CVD who sent at least one message between CVD and depression diagnoses (N = 2,314) took 1,746 days (4.78 years) from CVD diagnosis to depression diagnosis (**Figure 3**). The LLMs identified individuals with depression 660 days (1.8 years, Llama 3.1 8B) and 516 days (1.41 years, MedGemma 4B) earlier than formal diagnosis when applying the median frequency of symptom presentation, which was three, as a threshold to identify those with depression across 500 randomly sampled patients with CVD.

**Figure 3.**
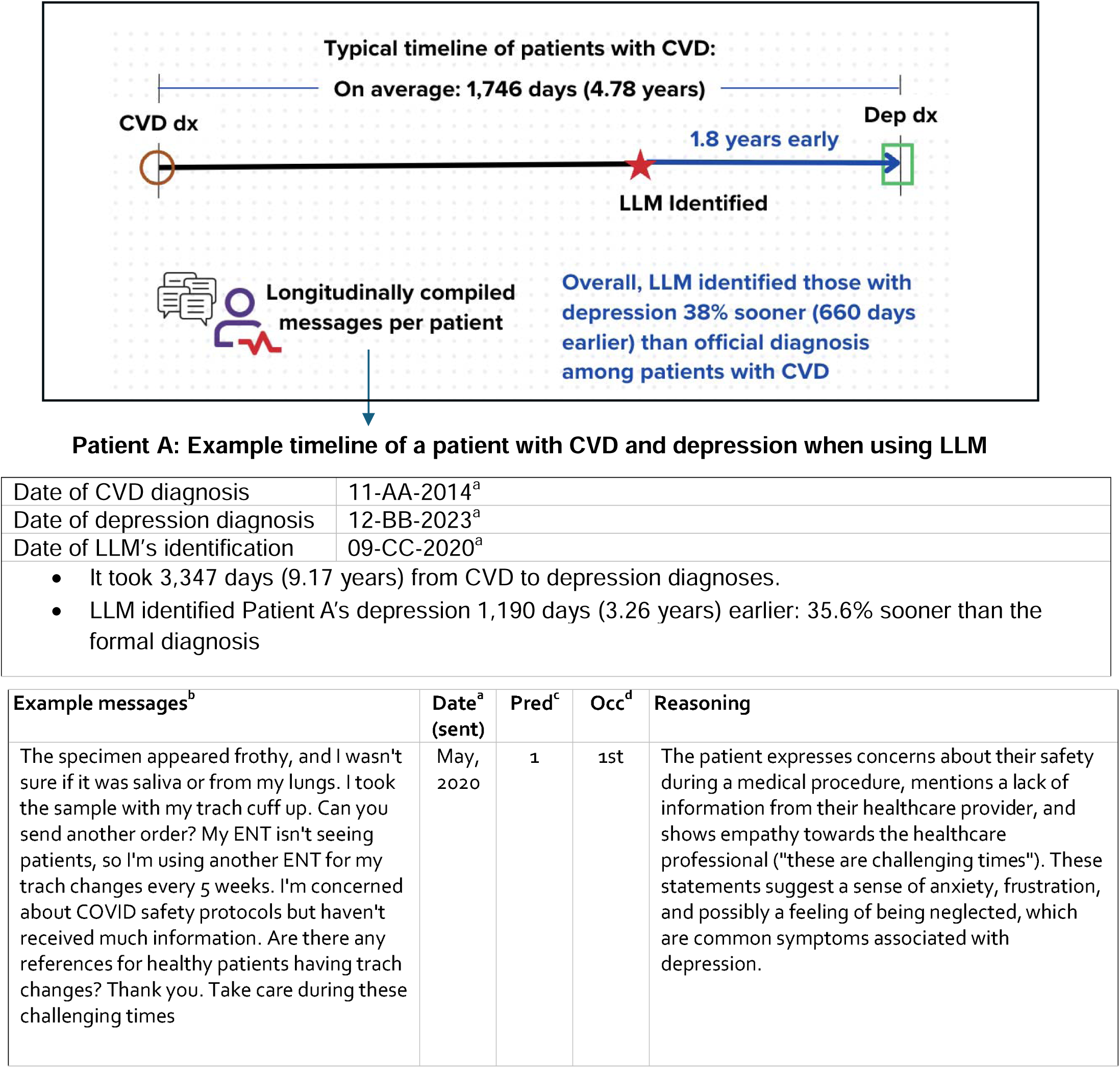
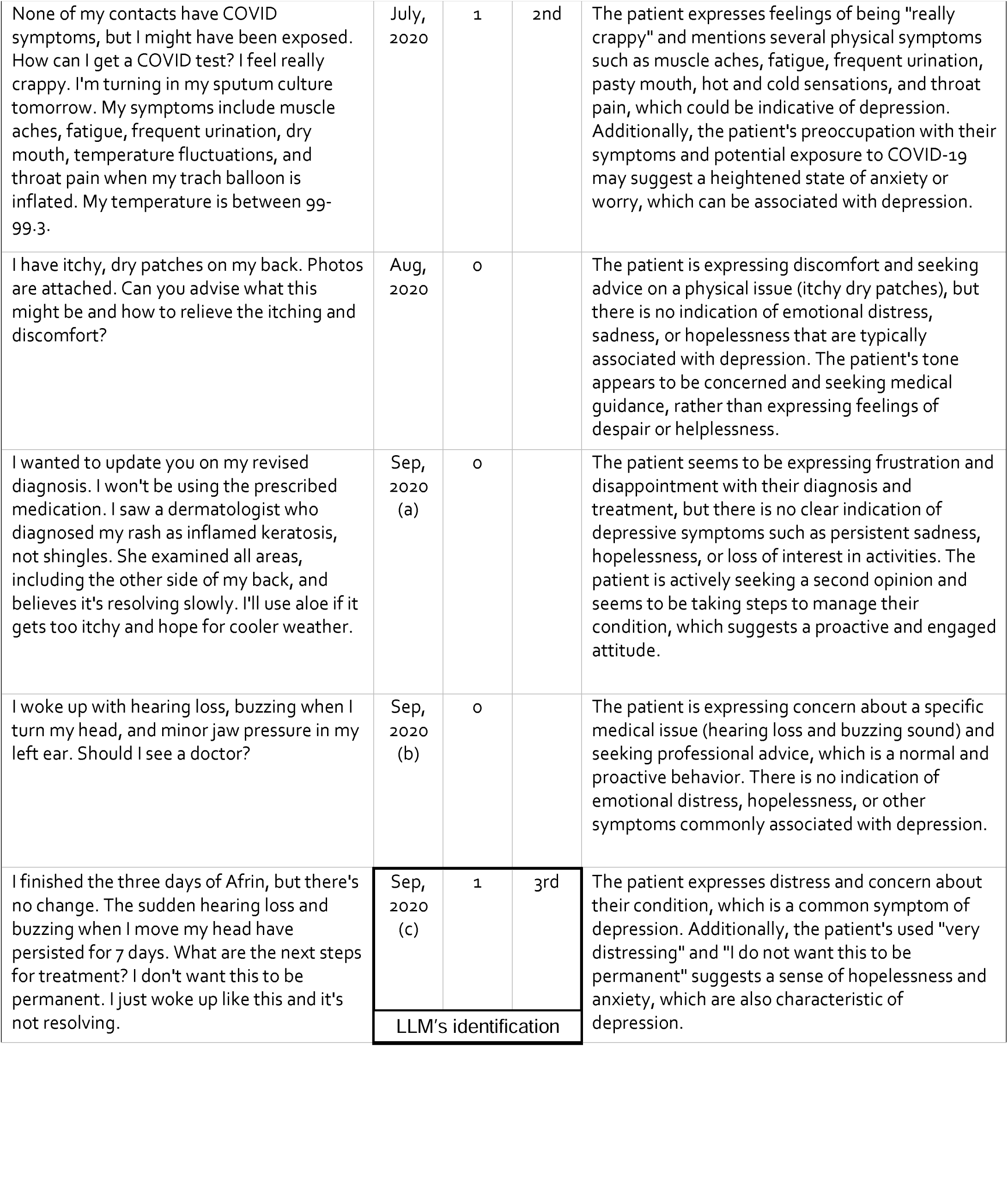
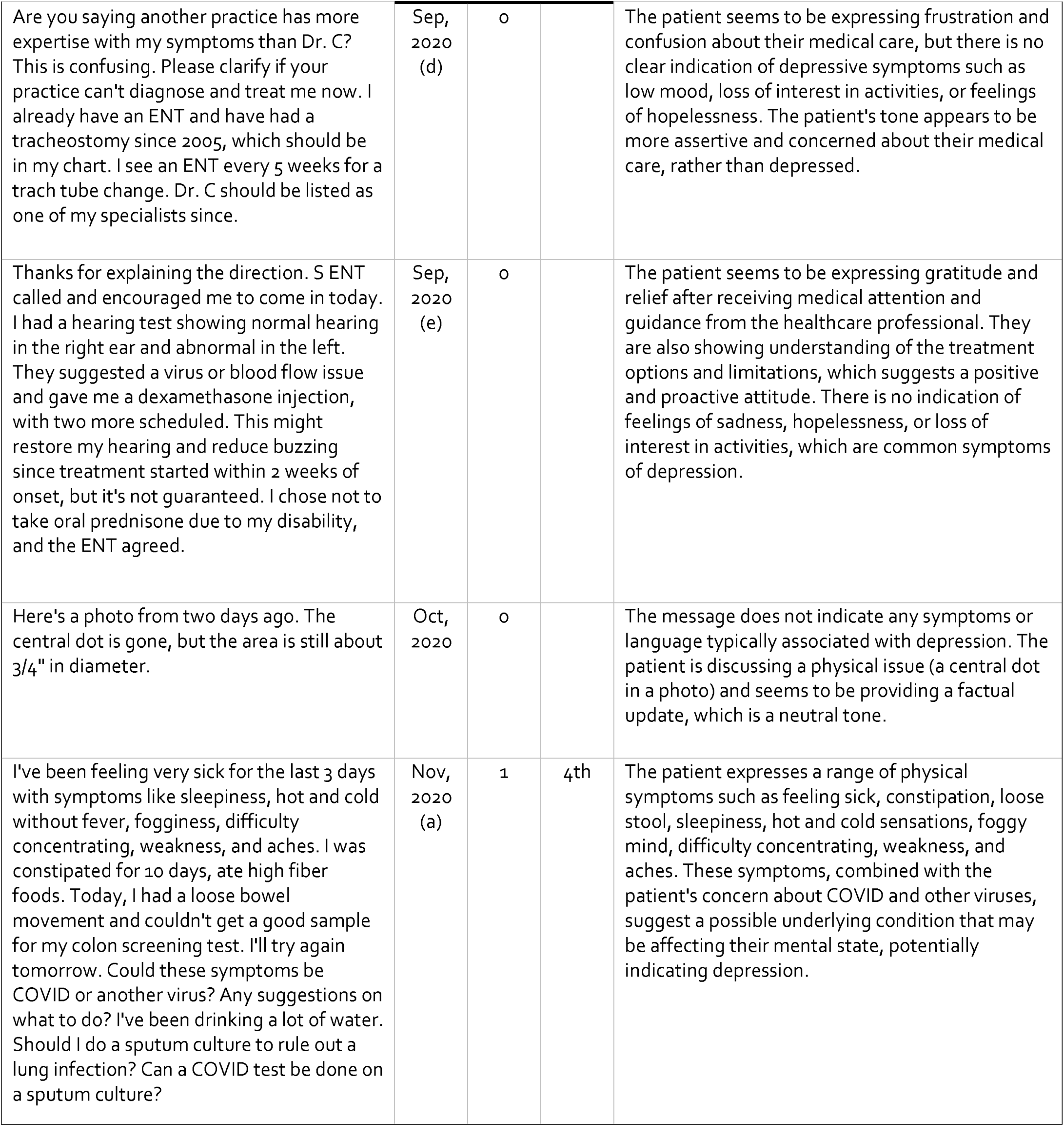
**LLM’s capacity to shorten the interval between CVD and depression diagnoses for early detection** a. We used the exact date and time format for calculation (Year-Month-Date, Hour-Minute-Second). In this Figure, we simplified them for privacy; b Patient messages in this Figure were slightly modified to preserve privacy and approved to share with the public. For analyses, we used the original forms of patient messages, which is available in eTable 1; c. Pred: Prediction was labeled by the LLM (“1” for presence of depression symptom and “0” for absence of depression symptom in the message); d. Occ: Occurrence refers to the frequency of depression symptoms. In this study, we used the 3rd occurrence as a threshold to identify individuals with depression; hence, the date of the 3^rd^ occurrence was marked as the date of LLM’s identification.

## Discussion

Our study demonstrates that LLMs can identify individuals with depression among patients with CVD significantly earlier than the official diagnosis. On average, the LLM detected depression 660 days earlier (38% sooner) than the first charted diagnosis, with an 83% sensitivity over a 1,746-day assessment period, which is a typical timeline from CVD to depression diagnoses in our cohort. The LLM accurately pinpointed individuals with depression based solely on symptoms detected from long text messages, longitudinally combined over nearly five years, without any additional patient information, showing 51% PPV, which is comparable to the performance of Patient Health Questionnaire-9 (PHQ-9).^22^ This advances our understanding of LLMs’ capacity from data-level symptom classification to patient-level risk stratification.^23^ The findings underscore the potential of secure patient messages as a valuable data source and LLMs as effective tools for the early identification of depression among those with CVD. This approach not only facilitates timely diagnosis for depression but also holds promise for the early detection of essential symptoms in other chronic diseases, primarily diagnosed through symptoms.

The strong sensitivity demonstrated by small open-source LLMs in screening longitudinally combined messages is particularly impressive. This is noteworthy given that these messages were often lengthy—averaging 25 compiled messages—and lacked a logical flow, as they encompassed a random assortment of topics from different time points. We focused exclusively on patient-initiated questions and responses, excluding those from healthcare professionals.

This context-constrained text data from one-way communications might limit the LLMs’ ability to fully understand the patients’ issues. Despite these challenges, the LLMs evaluated each message by referencing specific words or sentences to precisely support their decisions rather than offering generalized answers, which aligned with previous findings from an LLM-based conversational agent for mental health therapy.^24^ They demonstrated a nuanced understanding of depression, going beyond simply identifying terms like ’depressed’ or ’anxious.’ Instead, they considered multiple aspects of each patient, drawing from the entire message context and making observations and evaluations.

In many cases, patients with CVD may experience depression-like physical symptoms, such as lack of energy or sleep problems, without recognizing them as signs of depression, leading to delayed diagnoses.^2,4^ Particularly, cardiopulmonary symptoms were one of the most reported physical symptoms among individuals with anxiety-depressive disorders, highlighting the importance of a non-self-screening based approach.^25^ LLMs with high sensitivity have significant potential in identifying these symptoms early through routine patient message screening by overcoming the patient-level barriers to timely symptom detection (e.g. the lack of symptom awareness and the lack of motivation for self-screening).^2,3^ This capability could potentially expedite proper diagnosis, potentially opening opportunities for timely care for patients who might otherwise face long delays.

With over 51% of PPV, comparable to the performance of PHQ-9 —a reliable and valid measure for screening, diagnosing, and monitoring major depression —LLMs demonstrate strong potential as an effective screening agent in real-world settings. Additionally, our findings indicate that implementing a threshold can improve the model’s PPV (up to 12%) and sensitivity (up to 51%), providing valuable insights into an empirical cut-off for mental health screening.

While the frequency of depression symptoms presented in patient messages between CVD and depression diagnoses varied widely among individuals (ranging from 1 to 1,017), applying the median frequency as a threshold significantly improved the performance measurements, compared to the baseline performance without a threshold. LLM-assisted symptom screening and detection can help address clinician-level barriers to timely depression diagnosis, such as limited clinician availability.^11,26^ This approach could alert primary care providers (PCPs) for professional assessments. However, the workflow needs to be carefully designed to avoid additional clinician burden. Through a well-thought-out workflow, by assisting PCPs in focusing on patients at risk, this approach could reduce missed diagnoses and inappropriate referrals. It ensures that individuals who need a proper diagnosis receive timely care, addressing key bottlenecks such as misallocated patient triage, and long wait times that contribute to delays in the diagnostic process.^27^

The ability of LLMs to identify depression 1.8 years earlier than the first charted diagnosis, using only the symptoms described by patients without additional medical records, is remarkable.

However, interpretation requires caution, as the reference point for depression diagnosis in this study may not precisely reflect the onset of depression due to the disorder’s gradual development and varying symptoms over time.^28^ We interpret these results as evidence that LLMs can effectively pinpoint individuals at risk of depression before their formal diagnosis. A previous study reported that the median time to diagnosis of mood disorder if untreated within a year from symptom onset was approximately 5 years, which aligned with our findings (average 4.78 years).^29^ Detecting symptoms 1.8 years in advance provides a valuable window for timely intervention, given that there is another delay in proper evaluation.^27^ To address further downstream barriers to timely treatment and care after identifying those with depression through LLM-assisted screening barriers (e.g. shortage of available psychiatrists or mental health clinicians), a multi-disciplinary approach could be an option, perhaps involving digital mental health programs for eligible individuals.^30–33^ The emerging digital mental health intervention has demonstrated benefits in clinical effectiveness, including those with chronic disease or older adults, particularly during the initial stages of intervention, though potential risks should be monitored.^34–36^ Future directions may include exploring workflows that integrate LLM-assisted symptom screening with professional assessments for diagnosis, followed by tailored interventions based on severity, potentially leveraging available digital mental health solutions.^37–39^ Additionally, conducting an economic evaluation of AI assistance will be crucial to quantify its benefits relative to costs within the current healthcare system, assessing the scalability and sustainability of this innovative screening approach in real-world settings.^40^ Finally, incorporating patients’ perspectives into the integration of AI assistance in mental health care can help ensure the ethical use of AI tools in healthcare.^39,41^

This study has limitations. First, we focused solely on co-morbid depression among patients with CVD while not accounting for the many other potential psychiatric (e.g. generalized anxiety disorder or obsessive-compulsive disorder) and medical (e.g. diabetes or obesity) comorbidities in these patient populations.^42,43^ Further subgroup analysis of LLM performance is warranted to identify which patients would benefit the most from AI-assisted screening. Second, we are limited by our dataset, including the use of ICD-10 codes as the reference standard for diagnosis, the use of EHR data from a single institution, and the limited timeline of available data (2014-2024).^44,45^ Third, we employed small LLMs in a zero-shot setting due to their strong performance in text-based symptom detection. However, exploring approaches such as fine-tuning or few-shot learning could be beneficial to maximize the flexibility of open-source models, which could further enhance their performance. Lastly, about 50% of participants were older adults (75+), with messages often sent from caretakers. Although this could potentially dilute the representativeness of patient groups, the LLMs effectively distinguished patient symptoms from caretaker input.

## Conclusion

LLMs identified individuals with depression significantly earlier than the officially charted dates among patients with CVD, relying solely on longitudinal patient portal messages without additional medical information. By setting a threshold of three symptom presentations—the median frequency of depression symptoms prior to diagnosis—LLMs were able to identify patients at risk for depression with over 83% sensitivity while maintaining similar PPV to the PHQ-9 in patients with CVD. Among patients with an eventual diagnosis of depression, this early warning system may help overcome existing barriers to timely diagnosis and intervention. This novel application of LLMs used to screen patient message data could potentially revolutionize the care of chronic illnesses, not limited to depression, by facilitating automated symptom-driven diagnoses and interventions.

## Data Availability

The data (patient messages) used in this study is not publicly sharable.

## Author Contributions

Kim and Linos had full access to all the data in this study and take responsibility for the integrity of the data and the accuracy of the data analysis.

- Concept and design: Kim, Ma
- Acquisition, analysis, or interpretation of data: Kim, Torous, Adler-Milstein, van Roessel, Ma
- Drafting of the manuscript: Kim, Torous, and Rodriguez (CIR), Ma
- Critical review of the manuscript for important intellectual content: Kim, Torous, Adler-Milstein, van Roessel, Rodriguez (FR), Sharp, Pfeffer, Chen, Ma, Rodriguez (CIR), and Linos
- Statistical analysis: Kim, Ma
- Administrative, technical, or material support: Sharp, Pfeffer, and Linos
- Supervision: Rodriguez (CIR) and Linos

